# Physical Activity and Type 2 Diabetes Risk: Heterogeneity in the Dose-Response by Obesity and Age

**DOI:** 10.1101/2025.10.19.25338324

**Authors:** Jing Liu, Qingtao Zeng

**Affiliations:** Physical Education College of Henan University, Kaifeng 475001, Asia, China

**Author notes:** Correspondence: Qingtao Zeng.

**Keywords:** Physical Activity, Type 2 Diabetes, Effect Modification, Obesity, Age Factors

## Abstract

**Background:** Clinical evidence increasingly identifies individuals with severe mental disorders as a population exhibiting markedly elevated susceptibility to diabetes and its associated complications. Within this context, physical inactivity and prolonged sedentary behavior emerge as prominent, modifiable risk factors contributing to type 2 diabetes mellitus (T2DM). However, a critical gap persists regarding whether obesity and age significantly modify the protective association of physical activity (PA) and T2DM, and the precise nature of this dose-response relationship remains inadequately characterized.

**Methods:** We analyzed cross-sectional data from 5,042 adult participants in the 2017-2020 National Health and Nutrition Examination Survey (NHANES). We applied weighted multivariable logistic regression models to evaluate the PA-T2DM association. Furthermore, to delineate the dose-response relationship, we utilized restricted cubic splines (RCS), specifically testing for potential effect modification by body mass index (BMI) and age.

**Results:** After comprehensive adjustment for confounders, our analysis indicated that higher PA levels were independently correlated with a reduced T2DM risk. RCS dose-response analysis revealed nuanced effect modification patterns. Specifically, PA’s protective effect demonstrated a clear "BMI gradient"—most pronounced in normal-weight individuals, somewhat attenuated in the overweight group, and absent in participants with obesity. Concurrently, an "age stratification" pattern emerged; the association was strongest in middle-aged adults, exhibited a borderline significant trend among younger adults, and showed no significant association in older adults.

**Conclusion:** Our findings suggest that the protective effect of PA against T2DM is modified by obesity and age. Consequently, future preventive strategies should evolve from universal PA recommendations toward more precise, stratified guidance that incorporates individual BMI and age profiles. This approach appears particularly relevant for integrated care models targeting high-risk populations, such as individuals with mental disorders.

## 1 Introduction

The global burden of diabetes continues to escalate. By 2025, this epidemic presents a critical public health challenge that demands urgent attention. This epidemic exhibits two particularly concerning characteristics: a persistently high prevalence coupled with a disturbing trend toward earlier onset. In addition to immediate metabolic dysregulation, diabetes initiates a cascade of serious systemic complications,—notably cardiovascular and renal pathologies,—which substantially contribute to premature mortality. Recent estimates from the International Diabetes Federation (IDF) Atlas (11th edition) indicate an unprecedented surge in global diabetes cases, affecting approximately 589 million individuals worldwide^[1]^.

The diabetes landscape is further complicated by a pronounced increase in early-onset cases (diagnosis age <40 years), now affecting up to 8% of young adults in certain populations^[2]^. This shift is strongly correlated with a concurrent increase in obesity, unhealthy diets, and sedentary lifestyles. Management failures exacerbate this problem. A mere 21% of patients globally meet all recommended targets for blood glucose, pressure, and lipids. Concurrently, the high rate of undiagnosed cases (44.2%) means that a large population accumulates irreversible damage, significantly elevating the lifetime risk of cardiovascular and renal diseases^[3]^.

In recent years, numerous studies have indicated that obesity is a key risk factor for the development of type 2 diabetes^[4]^. An elevated BMI showed a significant positive correlation with the risk of developing type 2 diabetes. Extensive research has thoroughly explored and confirmed that physical activity is a key modifiable factor in the prevention and management of T2DM. Regular physical activity exerts a positive effect on glycemic control through multiple mechanisms, including improving insulin sensitivity, promoting glucose uptake and utilization, reducing visceral fat, and regulating inflammatory responses^[5]^. Different types of physical activity, such as moderate-to-vigorous physical activity (MVPA), aerobic exercise, and resistance training, have all been shown to confer varying degrees of benefit in reducing T2DM risk and improving patient outcomes.

A large-scale study based on the UK Biobank revealed that moderate-to-vigorous physical activity (MVPA) plays a significant mediating role in reducing mortality in diabetic patients. Furthermore, replacing sedentary behavior with MVPA can yield substantial gains in life expectancy for patients^[6]^.

Additionally, growing evidence suggests the need to develop personalized strategies for different patient subgroups. For instance, among patients with prolonged sedentary time, activity at any intensity helps reduce all-cause mortality risk, whereas for those with less sedentary time, light-intensity activity might suffice. This underscores the need to accurately quantify the interaction between activity levels and sedentary behavior^[7]^.

A critical evidence gap persists regarding special comorbid populations, particularly individuals with psychiatric disorders. The existing literature fails to provide a sufficiently granular analysis of the PA-T2DM risk relationship across key demographic strata—notably BMI and age subgroups. To address this limitation, we analyzed the NHANES 2017-2020 data to examine the independent and joint effects of modifiable behavioral factors. Using RCS modeling, we specifically assessed the effect modification by BMI and age, characterized the precise PA-T2DM risk relationship, and disentangled independent versus synergistic effects. These findings will inform targeted behavioral interventions for high-T2DM-risk populations, including those with psychiatric conditions.

## 2 Methods

### 2.1 Sample

As a population-based cross-sectional survey, the NHANES systematically collects comprehensive data on health and nutritional status from the non-institutionalized US household population^[8]^. Our analysis focused on a representative subsample of 5,042 adults aged ≥20 years from the 2017-2020 survey cycle. This four-year cycle encompassed approximately 15,000 households, with all participants meeting the mandatory two-year US residency requirement. The Research Ethics Review Board Of the National Center for Health Statistics (NCHS) Research Ethics Review Board approved the study protocol, and all participants provided written informed consent^[9]^.

A total of 15,560 participants were initially enrolled. We first excluded individuals aged <20 years and those with missing data on body mass index (BMI), poverty income ratio (PIR), alcohol consumption, smoking status, sleep duration, sedentary time, and physical activity, resulting in the exclusion of 10,478 participants and leaving 5,082 eligible individuals. After further excluding 40 participants with missing diabetes status data, a final total of 5,042 participants were included in the analysis.The following figure shows detailed information on the study design, sampling methods, and exclusion criteria (see Figure 1).

### 2.2 Measurement

#### 2.2.1 Covariates

Multiple covariates were considered in our analysis: sex, age, race/ethnicity, educational level, marital status, PIR, BMI, sleep duration, and sedentary time. Following WHO standards, we classified participants into three weight categories: normal weight (18.5–24.9 kg/m²), overweight (25.0–29.9 kg/m²), and obese (≥30.0 kg/m²)^[10]^. It is important to note that individuals with BMI <18.5 kg/m² were excluded from subgroup analyses due to the limited sample size in the underweight category—a methodological constraint that should be considered in interpreting the results. To capture potential effect modification across the life course, we adopted a life-course epidemiological framework^[11]^, stratifying participants into young (18–39 years), middle-aged (40–59 years), and older adults (≥60 years). Sedentary behavior was objectively measured using accelerometers, typically worn at the waist or wrist^[12]^. Smoking status was derived from self-report and categorized following the conventional approach used in major surveys such as the NHANES^[13]^: never smokers (≤100 lifetime cigarettes), former smokers (>100 cigarettes but quit), and current smokers (>100 cigarettes and still smoking).

#### 2.2.2 Physical Activity Assessment

We evaluated PA using the Global Physical Activity Questionnaire (GPAQ), expressing total volume as metabolic equivalent minutes per week (MET-min/week). This standardized metric encapsulates the cumulative energy expenditure from all bodily movements, incorporating both moderate-to-vigorous intensity activities (MVPA) and structured exercise into daily living^[14]^. The international standardization of MET-min/week as a summary measure enables consistent cross-study comparisons of weekly activity volumes^[15]^. It is worth noting that while this approach facilitates comparability, it may oversimplify the complex interplay between activity intensity, duration, and metabolic health—a consideration relevant to interpreting our findings.

#### 2.2.3 Type 2 Diabetes

T2DM is a complex metabolic disorder characterized by progressive insulin resistance coupled with inadequate compensatory insulin secretion^[16]^. For case identification, we applied internationally standardized diagnostic thresholds: a fasting plasma glucose (FPG) ≥7.0 mmol/L, and/or a 2-hour oral glucose tolerance test (OGTT) value ≥11.1 mmol/L, and/or a glycated hemoglobin (HbA1c) level ≥6.5%^[17]^. This multi-criteria approach, although well-validated, likely enhances diagnostic sensitivity—though it may introduce some heterogeneity in the identified patient population, a factor worth considering when interpreting our results.

### 2.4 Data analysis methods

All statistical analyses were conducted using Zstats software (based on R 4.3.1). To ensure nationally representative estimates and accurate variance calculations for the US adult population, our analytical approach fully accounted for the complex survey design of the NHANES—incorporating sampling weights, stratification variables, and primary sampling units as recommended in official guidelines^[18]^. This methodological decision strengthens the generalizability of our findings, although we note that the inherent limitations of secondary data analysis necessarily constrain analytical flexibility.

#### 2.4.1 Analysis of Baseline Characteristics

Continuous variables following normal distributions are presented as weighted means ± standard errors (SE), while non-normally distributed measures are summarized as weighted medians with interquartile ranges (IQR). For categorical measures, we reported weighted percentages (%). To evaluate group differences, we employed survey-weighted linear regression models for continuous outcomes and survey-weighted Rao-Scott chi-square tests for categorical variables—a methodological choice that appropriately accounts for the complex sampling design. It should be noted that while these weighted approaches enhance population representativeness, they may reduce statistical power compared to simple random sampling analyses, a factor that warrants consideration when interpreting non-significant findings.

#### 2.4.2 Multivariable Logistic Regression

To evaluate the independent relationship between PA and T2DM risk, we established a series of weighted multivariable logistic regression models, implementing sequential adjustment for confounding factors. All associations are reported as odds ratios (OR) with corresponding 95% confidence intervals (CI). Our model-building strategy proceeds hierarchically as outlined below.

1. Model 1 (Crude Model): Adjusted for basic demographic factors: age (continuous), sex (male/female), and race/ethnicity.
2. Model 2 (Socioeconomic Model): Model 1 + socioeconomic factors: educational level and family income-to-poverty ratio (PIR, continuous).
3. Model 3 (Fully Adjusted Model): Model 2 + lifestyle factors and body mass index (BMI): smoking status, drinking status, and BMI (continuous).

#### 2.4.3 Dose-Response Analysis Using Restricted Cubic Splines

To explore the potential nonlinear relationship between PA volume (continuous independent variable) and T2DM risk (binary dependent variable) and to test for effect modification by BMI and age, we performed survey-weighted RCS regression with logistic models.

1. Knot placement: Four knots were placed at the 5th, 35th, 65th, and 95th percentiles of the PA distribution.
2. Reference Value: The reference value for the OR was set at the median PA level (2,760 MET-min/week).
3. Test for Nonlinearity: The likelihood ratio test was used to assess nonlinearity by comparing the model with only the linear term to the model containing both the linear and spline terms.
4. Stratified Analysis: Separate RCS models were fitted within strata defined by BMI (normal-weight, overweight, obesity) and age (young, middle-aged, and older) groups. Each stratified model was adjusted for all covariates listed in Model 3 (except when stratified by BMI).

## 3 Results

### 3.1 Demographic Characteristics

In this study, normally distributed continuous data were presented as Mean ± Standard Deviation (SD), and comparisons between two groups were performed using the independent samples t-test. Non-normally distributed continuous data are presented as Median (Q , Q ) and were compared using the Mann-Whitney U test. Categorical data are presented as n (%), and group comparisons were conducted using the chi-square test or Fisher’s exact test, as appropriate. A two-sided p-value < 0.05 was considered statistically significant for all group comparisons.

The study included 5,042 adults aged 20 years and older from the 2017-2020 NHANES survey cycles who provided complete data on physical activity, sleep duration, sedentary time, and other demographic information. Among the 5,042 participants, 902 (17.9%) were classified as having T2DM (yes) and 4,140 (82.1%) as non-diabetic (no). Participants in the ’Yes’ group were older (mean age 59.76 ± 13.05 years vs. 46.56 ± 17.04 years; *p* < 0.001) and had a higher BMI (32.78 ± 7.65 kg/m² vs. 29.16 ± 7.18 kg/m²; *p* < 0.001). The ’Yes’ group also had a lower median family income-to-poverty ratio (PIR) (2.26 [interquartile range (IQR) 1.19–4.19] vs. 2.46 [IQR 1.27–4.62]; *p* = 0.030) and a lower median physical activity level (1800.00 [720.00–5040.00] MET-min/week vs. 2880.00 [1040.00–8400.00] MET-min/week; *p* < 0.001). No statistically significant differences were observed in sleep duration (*p* = 0.215) and sedentary time (*p* = 0.927). For categorical variables, significant group differences were observed for sex (*p* = 0.002), race/ethnicity (*p* < 0.001), marital status (*p* < 0.001), educational level (*p* < 0.001), drinking status (*p* < 0.001), and smoking status (*p* < 0.001) (see Table ). Table 1 summarizes the baseline characteristics of the study participants stratified by diabetes status.

### 3.2 Initial Associations of Physical Activity, Sedentary Behavior, and T2DM

The results of the crude model (Model 1), adjusted for basic demographic variables, indicated significant associations between several factors and T2DM risk. Specifically, men had a significantly higher risk of T2DM than women (OR = 1.26, 95% CI: 1.09-1.46). Mexican Americans demonstrated a higher risk than non-Hispanic Whites (OR = 0.65, 95% CI: 0.51-0.83). Increasing age was significantly associated with greater risk (OR = 1.05 per year). Physical activity level showed a protective effect (OR = 0.99); however, it is important to note that this OR was very close to 1, and its clinical significance requires further evaluation in subsequent models. No significant association was found for sedentary time in this model (*p* = 0.806), suggesting that its effect might be confounded by other factors and requires further investigation in more fully adjusted models. These findings provide preliminary evidence for subsequent stratified analyses.

### 3.3 Association of Physical Activity with T2DM After Controlling for Socioeconomic Status and the Emergence of Sedentary Behavior Effects

Results from Model 2, which was adjusted for socioeconomic status, revealed significant changes in racial/ethnic associations with T2DM due to the confounding effects of economic factors. The association for Non-Hispanic Black individuals shifted from non-significant to a significant protective effect (OR changed from 0.91 to 0.75), indicating negative confounding, where their lower socioeconomic status had masked an intrinsic protective effect. Conversely, the protective effect observed for Non-Hispanic White individuals was substantially strengthened (OR changed from 0.65 to 0.40), indicating positive confounding, whereby their higher socioeconomic status had exaggerated the initially estimated protective effect.

Our analysis of behavioral determinants revealed several noteworthy patterns. PA maintained a consistent and, significant inverse association with T2DM risk (OR=0.99) after socioeconomic adjustment, supporting its status as an independent protective factor. Interestingly, after controlling for socioeconomic confounders, prolonged sedentary time emerged as a significant risk factor (OR=1.01), suggesting that previous studies may have underestimated its detrimental impact. Age and sex exhibited stable associations with T2DM risk in our modeling approach. Additionally, we observed a dose-response relationship between educational attainment and diabetes protection, with higher education levels demonstrating progressively stronger protective effects(see Table ). **Table 2 summarizes the odds ratios and 95% confidence intervals for physical activity and sedentary behavior across all three sequential adjustment models.**

### 3.4 Independent Association of Physical Activity with T2DM After Adjusting for BMI

In the fully adjusted model (Model 3), our findings revealed a persistent protective association between PA and T2DM risk independent of BMI (OR=0.99, *p* =0.039). Simultaneously, this model identified BMI as the most substantial independent risk factor (OR=1.09, *p* <0.001). A notable observation emerged regarding sedentary behavior: its previously significant association with T2DM risk in Model 2 was completely attenuated after BMI adjustment ( *p* =0.570). Furthermore, the associations with older age, male sex, and significant racial/ethnic disparities remained stable. This model clarifies the causal pathways among risk factors and underscores the independent value of physical activity in T2DM prevention, which is not entirely subsumed by its relationship with obesity(see Figure 2. Forest plot for the fully adjusted model). The fully adjusted model results are visualized in the forest plot Figure 2.

### 3.5 Association between Physical Activity and T2DM Stratified by BMI

To further investigate the potential modifying effects of weight status on the protective association of physical activity, we employed RCS curves to examine the nonlinear relationship between physical activity volume and the prevalence of T2DM.

In the normal-weight population, at lower activity levels (e.g., <10,000 MET-min/week), the risk of T2DM was significantly higher than that at the reference level (OR > 1). When physical activity increased to moderate to high levels (approximately 20,000 MET-min/week), the odds ratio (OR) approached 1.

In the overweight population, at lower activity levels, the odds ratio (OR) for T2DM risk was approximately 1.5. When physical activity increased to its highest level (approximately 40,000 MET-min/week), the risk OR decreased to approximately 0.5. Furthermore, the statistical test did not reveal any significant evidence of a nonlinear relationship.

In the obese population, the overall association was not statistically significant ( *p* = 0.699). The entire dose-response curve fluctuated slightly around the null line (OR = 1), and its 95% confidence interval broadly included the null value (OR = 1) across the entire range of physical activity volumes. Similarly, no significant nonlinear trend was observed in this population(see Figure ). Figure 3. Dose-response relationship between physical activity (MET-min/week) and type 2 diabetes risk stratified by body mass index categories.

### 3.6 Association between Physical Activity and T2DM Stratified by Age

To further investigate the potential modifying effects of age on the protective association of physical activity, we employed RCS curves to examine the nonlinear relationship between physical activity volume and the prevalence of T2DM.

In the young adult subgroup, the curve shape indicated a significantly elevated risk of T2DM (OR > 1) at low physical activity levels. As the activity volume increased, the odds ratio gradually decreased and approached 1. However, the wide 95% confidence interval across the entire curve, particularly in the high activity range, indicates substantial uncertainty in these risk estimates.

In the middle-aged adult subgroup, at lower physical activity levels (e.g., <10,000 MET-min/week), the risk of T2DM was significantly higher than that at the reference level (OR > 2.0). When physical activity increased to approximately 15,000 MET-min/week, the OR dropped rapidly to approximately 1.0, indicating that the risk returned to the population average level.

In the older adult subgroup, the confidence intervals for the odds ratios broadly encompassed 1.0 across almost all physical activity levels, indicating highly uncertain risk estimates that cannot be distinguished from a null association. Furthermore, the analysis did not reveal any significant evidence of a nonlinear relationship(see Figure ). Figure 4. Dose-response relationship between physical activity (MET-min/week) and type 2 diabetes risk stratified by age groups.

## 4 Discussion

Logistic regression analysis of the NHANES 2017-2020 data revealed a highly significant association between physical activity and T2DM, influenced by the covariates of age and BMI. The protective effect of physical activity demonstrated a distinct "BMI gradient," being strongest in normal-weight individuals, attenuated in those with overweight, and absent in those with obesity. Concurrently, "age stratification" was observed, with the protective effect being strongest in middle-aged adults, showing a borderline significant trend in young adults, and demonstrating no significant association in older adults. Consequently, we separately discuss "BMI, Physical Activity, and T2DM", "Age, Physical Activity, and T2DM" in the following sections.

### 4.1 BMI, Physical Activity, and T2DM

Existing evidence posits that BMI is a central mediator in the sedentary behavior-diabetes relationship^[19]^. Although weight management remains a fundamental goal, our analysis suggests that PA provides independent protective benefits beyond its role in BMI regulation. Notably, in normal-weight individuals, we observed an essentially linear dose-response relationship between PA and T2DM risk reduction^[20]^. This continuous pattern—devoid of apparent saturation thresholds—implies that even modest PA increments may yield measurable risk reductions in this population.

Our data revealed a striking BMI-dependent gradient in the PA’ protective efficacy. The association was most pronounced in normal-weight individuals ( *p* < 0.01), modestly attenuated but still significant in overweight participants ( *p* < 0.05), and completely absent in those with obesity ( *p* > 0.05). This progressive attenuation pattern aligns with emerging evidence that obesity’s detrimental metabolic effects may overwhelm PA’s protective mechanisms^[21]^. Most notably, the complete absence of a significant PA-T2DM association in the obesity subgroup raises important clinical questions regarding intervention prioritization.

These findings strongly suggest that for individuals with obesity, a singular focus on PA augmentation may yield limited benefits. Instead, combined strategies addressing both weight management and PA promotion appear to be essential, with our data suggesting a possible primacy of weight control. This consideration is particularly crucial for vulnerable subgroups such as individuals with severe mental disorders, especially those prescribed weight-promoting medications such as olanzapine. Future studies should validate these findings in dedicated mental health cohorts and develop integrated care models that address physical activity and mental health needs concurrently. Ultimately, our results advocate for a departure from uniform PA recommendations toward BMI-stratified precision prevention paradigms.

### 4.2 Age, Physical Activity, and T2DM

Age emerged as a consistent independent determinant of T2DM risk, with each additional year associated with a 5% increase in odds (OR=1.05). Among younger adults, PA showed a borderline protective trend, possibly reflecting lower baseline risk and limited statistical power^[22]^.

While previous studies have established cardiometabolic benefits of MVPA timing in obese populations with T2DM^[23]^ and explored minimal exercise thresholds for glycemic control^[24]^, they did not examine how such thresholds vary across adult lifespans.

Our age-stratified analyses revealed progressive attenuation of the PA-T2DM protective association with advancing age. Younger adults exhibited a modest linear dose-response relationship, suggesting that achieving approximately 20,000 MET-min/week may substantially reduce T2DM risk. Middle-aged adults demonstrated a robust nonlinear association, supporting the concept of a "golden period" for lifestyle interventions^[25]^. Consistent with life-course epidemiological frameworks^[26]^. Notably, a critical threshold of approximately 15,000 MET-min/week was observed, with maximal benefit achieved between 20,000-25,000 MET-min/week.

In contrast, no significant PA-T2DM association was observed in older adults, likely reflecting confounding by multimorbidity and the healthy survivor effect^[27]^. Age-related physiological changes, such as reduced fast-twitch muscle fiber composition^[28]^ may simultaneously diminish metabolic efficiency and physical capacity, obscuring true PA effects.

Our age-stratified findings align with a growing body of NHANES-based research demonstrating that the relationship between modifiable lifestyle factors and health outcomes varies significantly across age groups. For instance, studies have identified nonlinear dose-response associations between dietary niacin intake and osteoporosis risk in older adults^[29]^, between riboflavin intake and osteoporosis risk in adult women^[30]^, and between high-fat diet and reduced bone mineral density in older adults^[31]^. Additionally, low dietary choline intake^[32]^, dietary protein intake^[33]^, and hip fracture-related factors^[34]^ have all been shown to exhibit age-dependent effects on bone health. Collectively, these studies underscore that age serves as a critical effect modifier across diverse health outcomes, reinforcing the importance of age-stratified approaches in both epidemiological research and clinical practice.

Collectively, these findings suggest that middle adulthood may represent the optimal window for implementing lifestyle interventions targeting T2DM risk reduction in vulnerable populations, including those with mental disorders.

### 5 Research limitations and future perspectives

This study employed a cross-sectional design, making it difficult to fully establish causal relationships. Issues such as "interference from comorbidities" and the "healthy survivor effect" represent typical confounding problems owing to reverse causality^[35]^. In older age (the period of effect attenuation), the independent statistical association of the protective effect disappeared. This does not imply that physical activity is unimportant for older adults; rather, its effect is masked by stronger factors (comorbidities and, survivor bias). These comorbidities significantly increase health risks (directly increasing the odds ratio) and severely restrict an individual’s physical activity capacity (directly reducing physical activity levels) ^[36]^.

These methodological challenges introduce substantial confounding factors when interpreting the PA-health outcomes relationship^[37]^. Rather than representing a direct causal pathway, the observed "physical inactivity-high risk" association appears largely attributable to comorbid conditions that simultaneously limit activity capacity and increase disease susceptibility. This confounding structure may mask the subtle protective effects of PA that remain undetectable within conventional observational frameworks.

Our analysis further suggests that older adults with high PA levels likely constitute a select population with inherent health advantages^[38]^. Their preserved functional capacity appears to reflect lifelong health behaviors, genetic predispositions, and socioeconomic resources, both of which enable sustained activity and directly promote health. This pattern supports a "health-determines-activity" paradigm rather than presuming uniform benefits from activity interventions across heterogeneous aging populations. Conversely, we propose that low activity levels in advanced age frequently indicate pre-existing multimorbidity rather than the primary causing it^[39]^.

Additionally, we must consider residual confounding from mental health conditions such as depression, which may simultaneously reduce PA and directly elevate diabetes risk through neuroendocrine pathways^[40]^. Building on the current findings, future investigations should employ genetically informed designs—such as Mendelian randomization—and incorporate comprehensive, deep-phenotyping frameworks exemplified by large-scale cohorts such as the German National Cohort^[41]^. Such an approach is critical to better disentangle these complex relationships. Furthermore, longitudinal studies featuring repeated measures of PA, metabolic parameters, and psychological factors are needed to clarify the temporal ordering of these associations and identify optimal intervention targets across the life course.

Beyond addressing methodological limitations, our findings suggest several directions for clinical translation. Future intervention trials should adopt BMI- and age-stratified designs to enable targeted physical activity prescribing. The null effect in individuals with obesity calls for mechanistic studies exploring whether alternative activity modalities (e.g., high-intensity interval training) may overcome obesity-related metabolic barriers. The identification of middle adulthood as an optimal intervention window supports integrating physical activity promotion into routine primary care for this age group. Finally, given the vulnerability of individuals with severe mental disorders, future research should develop scalable interventions within integrated care models that address both mental and physical health needs concurrently.

## 6 Conclusion

Based on our findings, we draw three central conclusions:

1. The protective association between physical activity and type 2 diabetes is jointly and significantly modified by body weight status (BMI) and age, demonstrating a distinct "BMI gradient" and "age stratification." **These findings challenge the "one-size-fits-all" approach to physical activity recommendations and underscore the necessity of personalized prevention strategies.**
2. For individuals with obesity, behavioral interventions must explicitly prioritize weight management, as we found no significant independent protective effect of physical activity in this group. This suggests that, in the absence of concurrent weight management, physical activity alone may yield limited metabolic benefits—a critical consideration for clinical counseling and public health messaging.
3. Middle adulthood represents a critical window for physical activity intervention, where we observed the strongest and most significant risk reduction for type 2 diabetes. **This life stage may offer the optimal return on investment for lifestyle interventions, highlighting an actionable opportunity for targeted screening and early intervention programs.**

Collectively, these findings advocate for a paradigm shift from universal physical activity recommendations toward precise, stratified guidance that integrates an individual’s BMI and age profiles—an approach that holds promise for enhancing the efficiency and effectiveness of diabetes prevention efforts in both clinical and population health settings.

## figure captions

Figure 1. Flow chart of subject selection

Table 1. Baseline Characteristics of the Study Participants

Table 2. Associations between physical activity and sedentary behavior with type 2 diabetes across sequential adjustment models.

Figure 2. Forest plot for the fully adjusted model

Figure 3. Dose-response relationship between physical activity and type 2 diabetes risk stratified by body mass index categories. (a) normal-weight group; (b) overweight group; (c) obesity group.

Figure 4. Dose-response relationship between physical activity and type 2 diabetes risk stratified by age groups. (a) young adults (18–39 years); (b) middle-aged adults (40–59 years); (c) older adults (≥60 years).

## Data Availability

This study analyzed publicly available, de-identified data from the National Health and Nutrition Examination Survey (NHANES) cycles 2017-2020. The NHANES protocol was approved by the National Center for Health Statistics (NCHS) Research Ethics Review Board (ERB). All participants in the survey provided written informed consent. The ethical approval for data collection in these cycles was covered under ERB Protocol #2011-17.

https://wwwn.cdc.gov/nchs/nhanes/Default.aspx

## Declarations

## Acknowledgements

We thank all the participants of this study.

## Data availability statement

All original contributions from this study have been included in the article; for further inquiries, please contact the corresponding author.

## Consent for publication

Not applicable.

## Clinical trial number

Not applicable.

## Funding

There is no funding or incentive to carry out this research.

## Author contribution

JL conceived and designed the study; JL and Q-TZ coordinated the study; JL collected the data; JL and Q-TZ analyzed and interpreted the data; JL drafted the manuscript; and Q-TZ revised the manuscript. All authors have read and approved the final version of the manuscript.

## Ethics approval and consent to participate

Human subject research was authorized by the National Center for Health Statistics. The studies were conducted in strict compliance with the local legal stipulations and institutional guidelines, ensuring that written informed consent was obtained from all participants before their involvement in the research.

## Statement of competitive interests

The authors state that they have no conflict of interest.

## Statement on Translation Tool Usage

DeepSeek’s latest model (DeepSeek-V3) was utilized as an AI translation assistant.

The following abbreviations are used in this manuscript

BMI: Body Mass Index
CI: Confidence Interval
GPAQ: Global Physical Activity Questionnaire
IQR: Interquartile Range
MET: Metabolic Equivalent
MVPA: Moderate-to-Vigorous Physical Activity
NHANES: National Health and Nutrition Examination Survey
OR: Odds Ratio
PA: Physical Activity
PIR: Poverty Income Ratio
RCS: Restricted Cubic Spline
SE: Standard Error
T2DM: Type 2 Diabetes Mellitus.

## Notes

### Competing Interest Statement

The authors have declared no competing interest.

### Summary of Updates

Author information changed, two authors were removed and a new author was added.

